# Pre-existing population immunity and SARS-CoV-2 variant establishment and dominance dynamics in the United States: An ecological study

**DOI:** 10.1101/2022.07.01.22277157

**Authors:** Pierre O. Ankomah, Mark J. Siedner, Roby P. Bhattacharyya

**Affiliations:** Massachusetts General Hospital, Boston MA, USA; Broad Institute, Cambridge MA, USA

## Abstract

We conducted an ecological analysis of the dynamics of Delta and Omicron establishment and dominance in U.S. states. Omicron became the dominant circulating variant later in states with higher population-level immunity. By contrast, population immunity did not impact the rates of transition from prior variants to either Delta or Omicron.

## INTRODUCTION

The pandemic of severe acute respiratory syndrome coronavirus-2 (SARS-CoV-2) has been characterized by the emergence of variants with competitive advantages in transmissibility and/or the capacity to evade pre-existing immunity [1]. A quantitative framework to understand how new variants displace circulating strains and the impact of population immunity on these dynamics is critical to inform public health preparedness for future waves.

Two recent variants of concern, Delta and Omicron, both reached global dominance, irrespective of geographic variation in rates of vaccination and/or prior infection [2][3]. These variants, however, became dominant under relatively different circumstances of competition with previously circulating strains. Whereas the Delta variant was somewhat more immune evasive than its predecessor, the Alpha variant, the Omicron variant is considerably more immune evasive than Delta [1]. In the United States, SARS-CoV-2 population immunity has historically varied considerably by state due to differences in prior infection and vaccine uptake [4]. Yet, it remains unclear how this variability impacts the establishment and dominance of variants with different immune evasiveness characteristics.

We performed an ecological study using publicly available data on SARS-CoV-2 incidence, vaccination rates and variant fractions to evaluate the dynamics of Delta and Omicron establishment and transition to dominance in the US. We assessed the relationship between population immunity to SARS-CoV-2, and the rate, date, and timing of variant takeover at a state level. We hypothesized that if immune evasion was a major driver of variant dominance dynamics, then both variants would become dominant sooner and faster in states with higher levels of pre-existing immunity, with a greater effect during Omicron takeover.

## METHODS

### SARS-CoV-2 Variant Data Sources and Definitions

We accessed SARS-CoV-2 genomic sequence data submitted to the Global Initiative on Sharing Avian Influenza Data (GISAID) (https://www.gisaid.org/) from all 50 US states between January 1 and August 13, 2021, and between November 24, 2021 and February 8, 2022. These periods spanned the first detection of the Delta and Omicron variants respectively, to a time when each consistently represented >99% of all sequenced genomes. For each period, we extracted the proportion of the emerging variant among total SARS-CoV-2 sequences, computed as 7-day averages. We identified the initial week during which a variant was first sequenced and designated it as the week of variant emergence if at least one additional isolate of the variant was identified within the subsequent three weeks. This constraint was implemented to avoid misclassifying stochastic introductions of a variant without subsequent sustained transmission as the true emergence of a new variant, a phenomenon that can be observed on occasion with over-dispersed spread [5]. It affected seven states during Delta emergence, and no states during Omicron emergence.

### Modeling/Statistical Methods to Evaluate Variant Takeover

We fit asymmetric logistic growth curves to changes in variant proportion over time and obtained curve-fit estimates for (i) maximum slope; (ii) inflection point, i.e. the time at which variant proportion was 50%; and (iii) time at which variant proportion was 10% [6]. In line with epidemiological reality, the lower and upper asymptotes of the curves were fixed at 0 and 100 respectively. Using these data, we derived three outcome measures of variant takeover: (i) takeover rate, defined as the maximum slope of the logistic curve; (ii) calendar date of variant dominance, estimated as the date variant proportion exceeded 50%; and (iii) time from establishment to dominance, computed as the time taken for variant proportion to increase from 10% to 50%. We estimated the calendar date of dominance to mitigate against the effect of variation in fractions of sequenced cases in different states impacting the likelihood of variant discovery, which would in turn alter the observed time to dominance. 10% variant fraction was chosen to define variant establishment to minimize the effect of stochastic fluctuations at lower variant fractions [7].

### Population Immunity Data Sources and Definitions

Using data from the US Centers for Disease Control (https://covid.cdc.gov/), and assuming a two-week delay between exposure and attainment of immunity, we estimated statewide immunity to COVID-19 using four definitions: (i) proportion fully vaccinated; (ii) proportion boosted; (iii) proportion with a prior infection and (iv) proportion with either prior infection or fully vaccinated/boosted. We defined the proportion with prior infection as the number of reported cases divided by the state population. We utilized this metric given the availability of time-resolved data and limitations on accuracy of other estimates of infection prevalence. For primary analysis, we assessed vaccine-induced population immunity using the most effective vaccine series available during variant takeover, i.e. the fully vaccinated proportion during Delta takeover and the boosted proportion during Omicron takeover [8]. We approximated combined immunity from infection or vaccination by estimating the fraction of the population without immunity as w=(1-p)*(1-v), where p and v are the previously-infected and vaccinated proportions respectively, then computing the total proportion immune as 1-w [9].

For each measure of variant takeover, we fit linear regression models to estimate the relationship with estimates of population immunity across different states. Code for the analysis is available at github.com/pankomah/variant_immunity. Analytic datasets are collated in Supplementary File 1.

## RESULTS

We fit logistic curves to estimate the proportion of SARS-CoV-2 infections attributable to Delta or Omicron variants in each state during the transition from Alpha and other circulating variants to Delta, and from Delta to Omicron (Figure S1, Table S1). In Figure 1, we graphically depict the relationship between variant takeover and combined population-level immunity from vaccination and infection. There was no statistically significant relationship between variant takeover rates and immunity for Delta (R=0.13, p=0.37; Fig. 1a) or Omicron (R=-0.14, p=0.32; Fig. 1b). Takeover for Omicron occurred at later dates in states with more immune populations (R=0.52, p<0.001; Fig. 1d), with a similar, but not statistically significant trend for Delta (R=0.24, p=0.091; Fig. 1c). There was also a statistically significant difference in time from establishment (10%) to dominance (50%) for Omicron (R=0.3, p=0.036; Fig. 1f), occurring over a longer period in states with higher population immunity, but not for Delta (R=-0.07, p=0.64; Fig. 1e).

**Figure 1.**
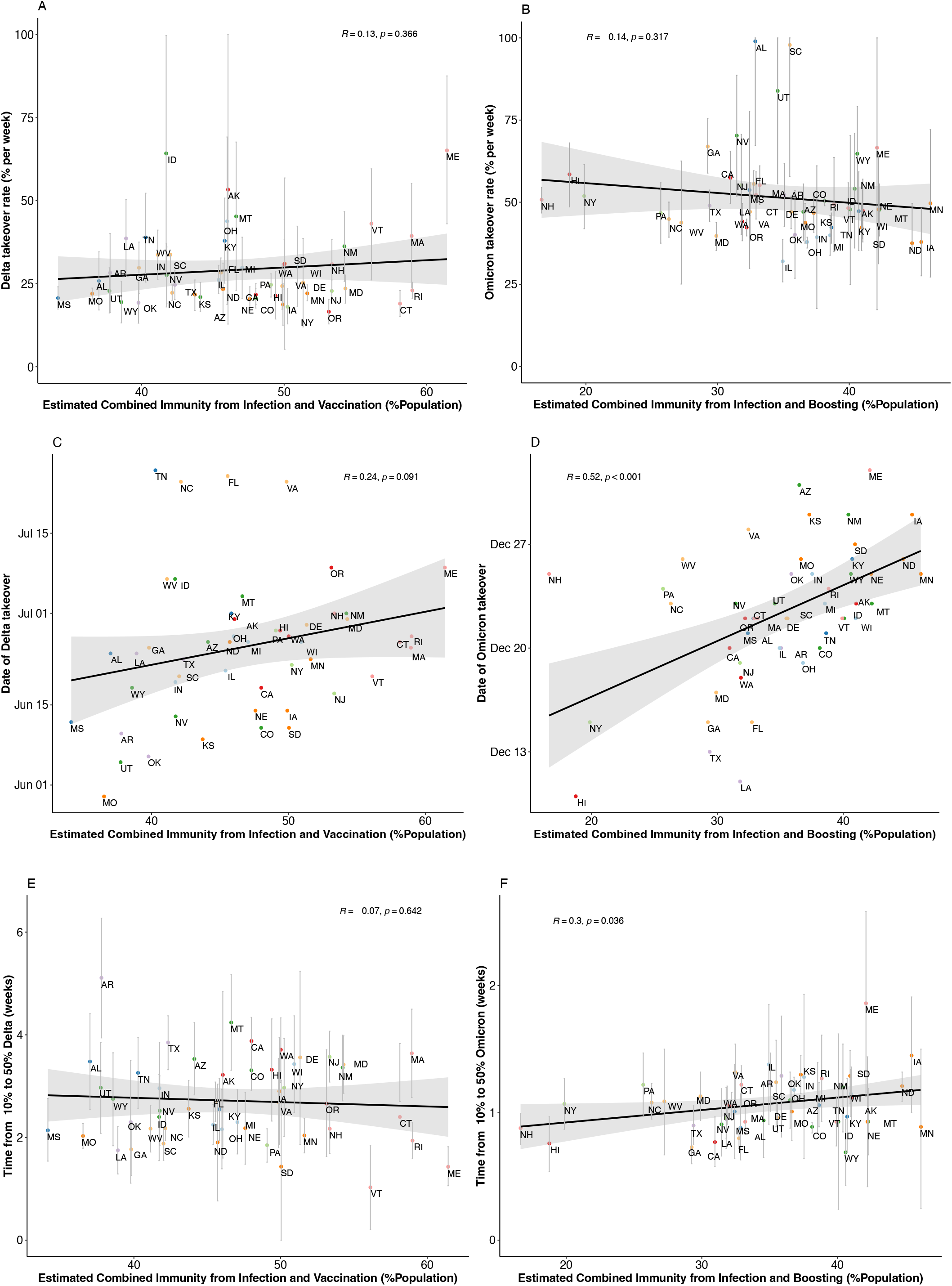
Delta and Omicron variant takeover and immunity in different US states. (A,B) Variant takeover rate in different states with 95% confidence intervals; (C,D) estimated initial calendar date at which a variant reached 50% of sequenced SARS-CoV-2 genomes in different states (date of takeover); (E,F) estimated time taken for variant proportion to increase from 10% to 50% of sequenced SARS-CoV-2 genomes in different states with 95% confidence intervals. States are identified by standard two-letter abbreviations; states in the same census geographic region are plotted with the same color. Error bars for takeover rates were limited to maxima and minima of 100 and 0 respectively. Left panel: Delta, Right panel: Omicron. Immunity is estimated by the combined proportion of the state’s population with identified SARS-CoV-2 infection prior to detection of the new variant in the state and either fully vaccinated (for Delta) or boosted (for Omicron) two weeks prior to variant proportion exceeding 50%. Pearson correlation coefficient (R) and p-value test results are shown for each plot.

In sensitivity analyses, we observed similar results when we limited estimates of population immunity to the most effective vaccination strategy for each variant, omitting recorded infections. There was no relationship between immunity and takeover rates, a statistically significant delay in Omicron takeover date (p<0.001) and time from establishment to dominance (p=0.009) in states with more boosted populations and a nonsignificant trend to delayed takeover date for Delta in states with higher proportions of fully vaccinated people (p=0.055) (Figure S2). When we used the fully vaccinated instead of the boosted proportion to represent immunity against Omicron (Figure S3) or documented preceding infection as the sole measure of immunity for both variants (Figure S4), no statistically significant relationships were observed between immunity and any takeover metric.

## DISCUSSION

In this ecological study, we tested the hypothesis that novel variants with greater immune-evasive capacity would become dominant faster in states with higher levels of population immunity. Contrary to our hypothesis, we found no statistically significant association between takeover rates of Delta or Omicron and state-level immunity. By contrast, we observed later takeover for Omicron in more immune states and a similar but not statistically significant trend for Delta.

These results suggest that population-level immunity did not enhance the rates at which the Delta or Omicron variants became dominant, despite greater immune escape capacity for each compared to major variants that were circulating at the time of their emergence. While preferential transmission among vaccinated individuals is expected for immune-evading variants [10], we speculate that this might have been counterbalanced by decreased rates of secondary transmission in these same immune sub-populations. For example, longitudinal cohort studies have demonstrated that full vaccination and boosting decrease the secondary attack rate among contacts of individuals infected with Delta or Omicron respectively and accelerate the rate of viral clearance, thereby decreasing potential subsequent transmission events [11]. The observation of a statistically significant delay in date of and time to Omicron but not Delta takeover, may be sequelae of the relatively greater ability of Omicron to infect immune sub-populations during its competitive circulation with Delta, moreso than for Delta with Alpha. As a result, Omicron’s dominance would be more likely slowed down by immunity compared to Delta.

Important caveats to our observations include limitations associated with use of publicly available data. For example, we used public case reporting to estimate immunity provided by prior infection, which likely substantially underestimates total infection rates, possibly to varying extents in each state. GISAID data on variant proportions is also a selected sample of all cases. In addition, unmeasured confounders such as public health mitigation policies, behaviors or population density may systematically vary by state and concurrently affect both vaccination rates and variant transmission. Finally, we do not account for the waning of vaccine-induced immunity in susceptibility to novel variant infections [12], or the mitigating impact of vaccination on disease severity or death.

Nevertheless, our results suggest that use of population-level data on vaccination and infection rates alongside assessments of changing variant proportion may provide a useful framework to understand how population immunity affects circulating SARS-CoV-2 variants. These findings do not support theoretical concerns about enhanced selection for immune-evasive variants as a drawback of widespread vaccination campaigns, since states with more measured immunity saw similar maximum takeover rates and similar or later time to dominance of emerging variants.

## Data Availability

All data produced in the present study are available upon request to the authors

## Notes

### Financial support

This work was supported by National Institutes of Health T32 AI007061 to P.A.

## Acknowledgments

We gratefully acknowledge the originating laboratories responsible for obtaining the specimens and the submitting laboratories where genetic sequence data were generated and shared via the GISAID Initiative.

## Potential conflicts of interest

None

## Figure Legends

**Figure S1.** Asymmetric logistic curve fits to weekly estimates of Delta (black circles) or Omicron (red triangles) as a proportion of all SARS-CoV-2 isolates in different states grouped by census geographic region.

**Figure S2.** Delta and Omicron variant takeover and vaccination-induced immunity in different US states. (A,B) Variant takeover rate in different states with 95% confidence intervals; (C,D) estimated initial calendar date at which a variant reached 50% of sequenced SARS-CoV-2 genomes in different states (date of takeover); (E,F) estimated time taken for variant proportion to increase from 10% to 50% of sequenced SARS-CoV-2 genomes in different states with 95% confidence intervals. States are identified by standard two-letter abbreviations; states in the same census geographic region are plotted with the same color. Error bars for takeover rates were limited to maxima and minima of 100 and 0 respectively. Left panel: Delta, Right panel: Omicron. Immunity is represented by the proportion of the state’s population fully vaccinated (for Delta) or boosted (for Omicron) two weeks prior to variant proportion exceeding 50%. Pearson correlation coefficient (R) and p-value test results are shown for each plot.

**Figure S3.** Omicron variant takeover and vaccination-induced immunity in different US states. (A) Omicron takeover rate in different states with 95% confidence intervals; (C,D) estimated initial calendar date at which Omicron reached 50% of sequenced SARS-CoV-2 genomes in different states (date of takeover); (E,F) estimated time taken for Omicron proportion to increase from 10% to 50% of sequenced SARS-CoV-2 genomes in different states with 95% confidence intervals. States are identified by standard two-letter abbreviations; states in the same census geographic region are plotted with the same color. Error bars for takeover rates were limited to maxima and minima of 100 and 0 respectively. Immunity is represented by the proportion of the state’s population fully vaccinated two weeks prior to variant proportion exceeding 50%. Pearson correlation coefficient (R) and p-value test results are shown for each plot.

**Figure S4.** Delta and Omicron variant takeover and infection-induced immunity in different US states. (A,B) Variant takeover rate in different states with 95% confidence intervals; (C,D) estimated initial calendar date at which a variant reached 50% of sequenced SARS-CoV-2 genomes in different states (date of takeover); (E,F) estimated time taken for variant proportion to increase from 10% to 50% of sequenced SARS-CoV-2 genomes in different states with 95% confidence intervals. States are identified by standard two-letter abbreviations; states in the same census geographic region are plotted with the same color. Error bars for takeover rates were limited to maxima and minima of 100 and 0 respectively. Left panel: Delta, Right panel: Omicron. Immunity is represented by the proportion of the state’s population with identified SARS-CoV-2 infection two weeks prior to detection of the variant in the state. Pearson correlation coefficient (R) and p-value test results are shown for each plot.

## Notes

### Competing Interest Statement

The authors have declared no competing interest.

### Funding Statement

This study did not receive any funding

