## Supplementary figures and images for "Pre-existing population immunity and SARS-CoV-2 variant establishment and dominance dynamics in the United States: An ecological study"

### Supplemental Figure 1

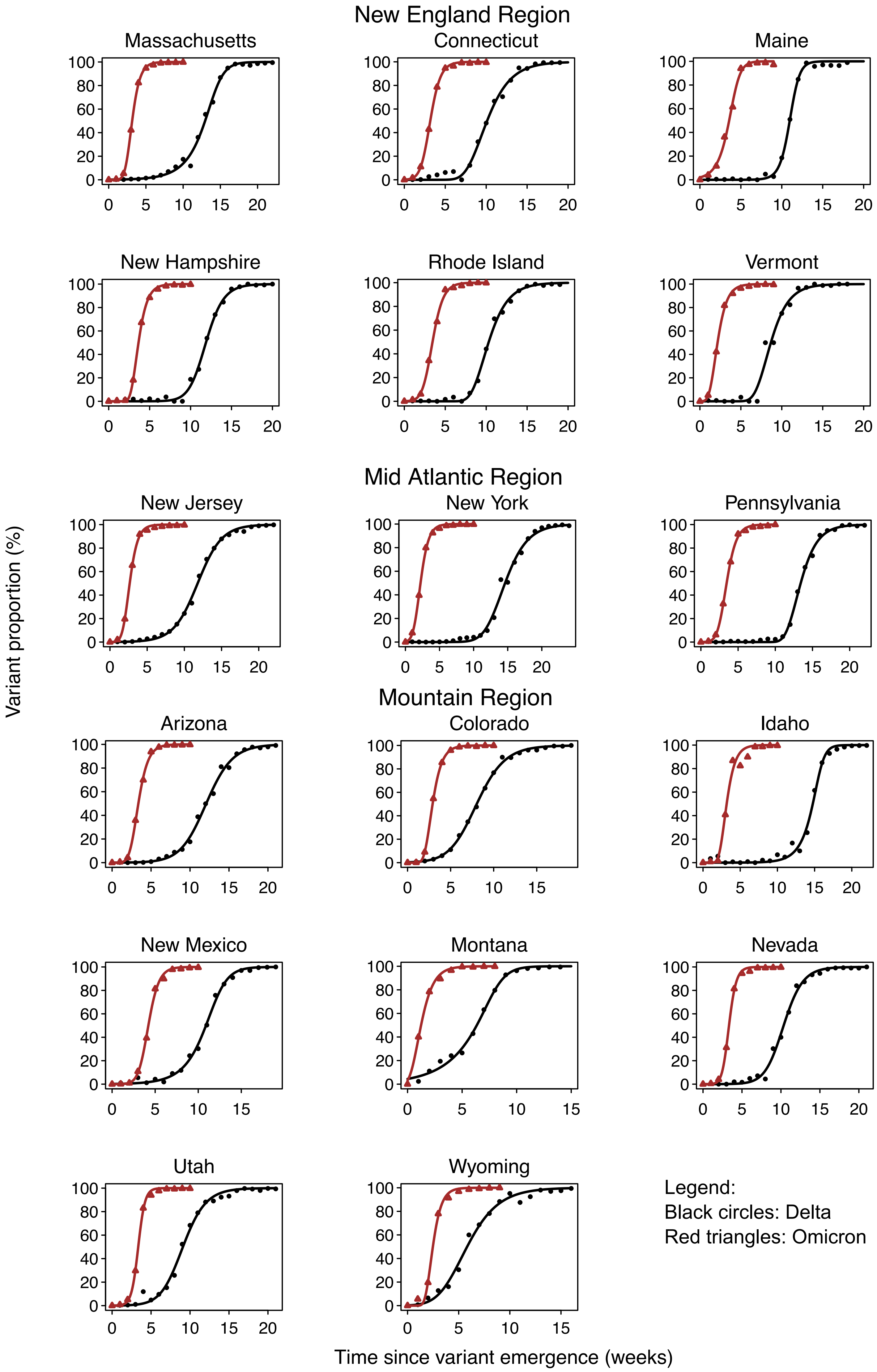

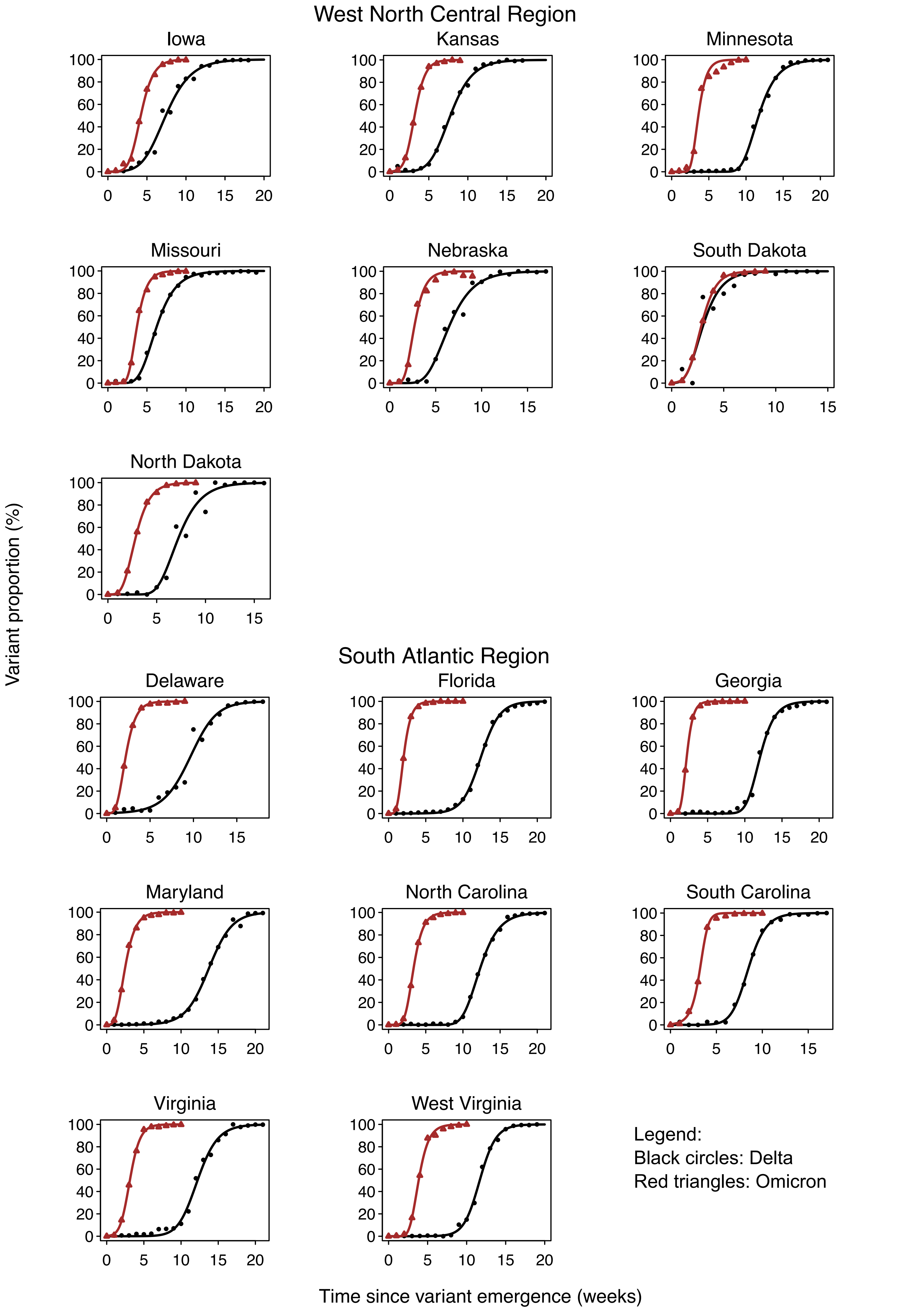

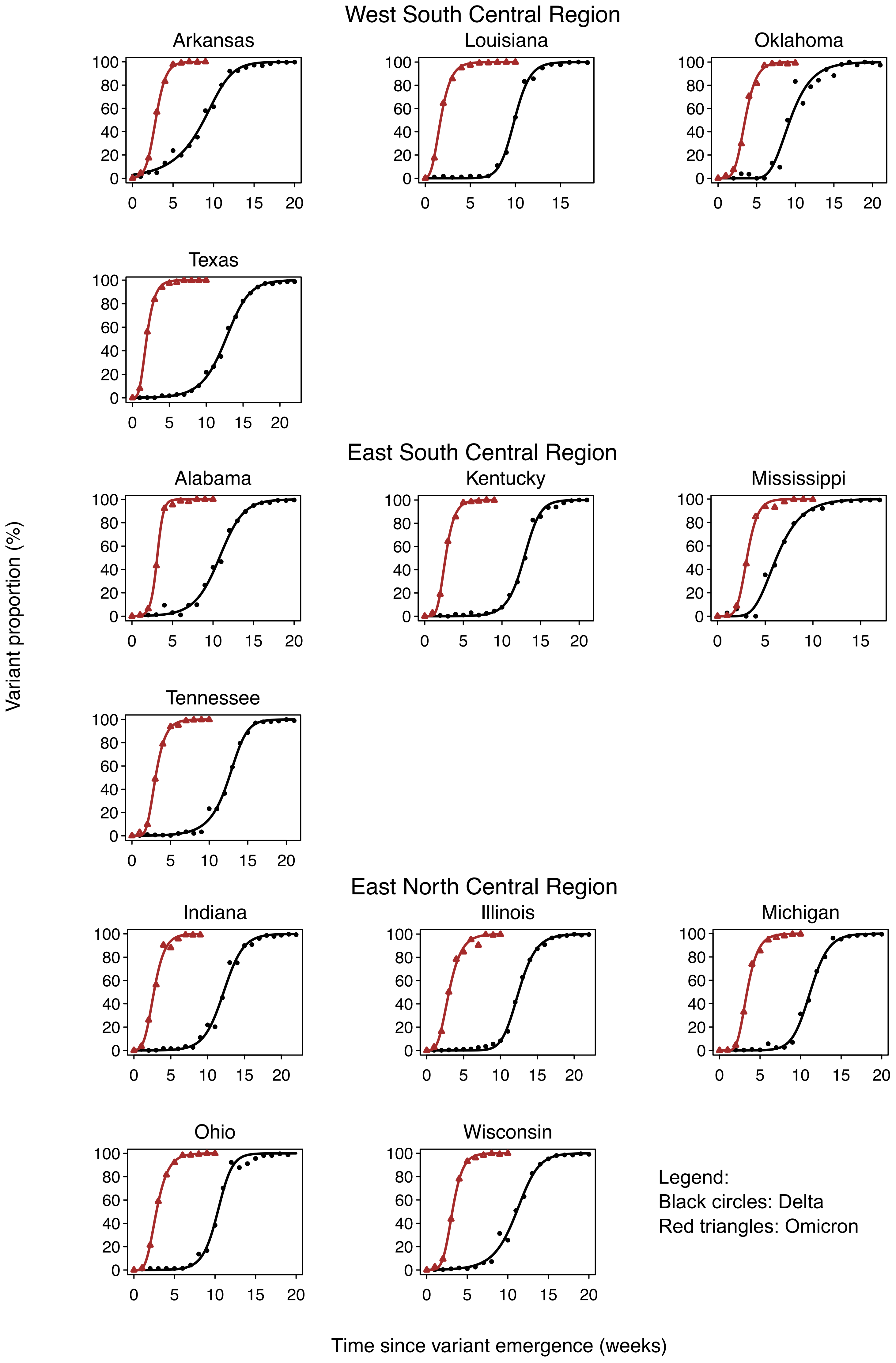

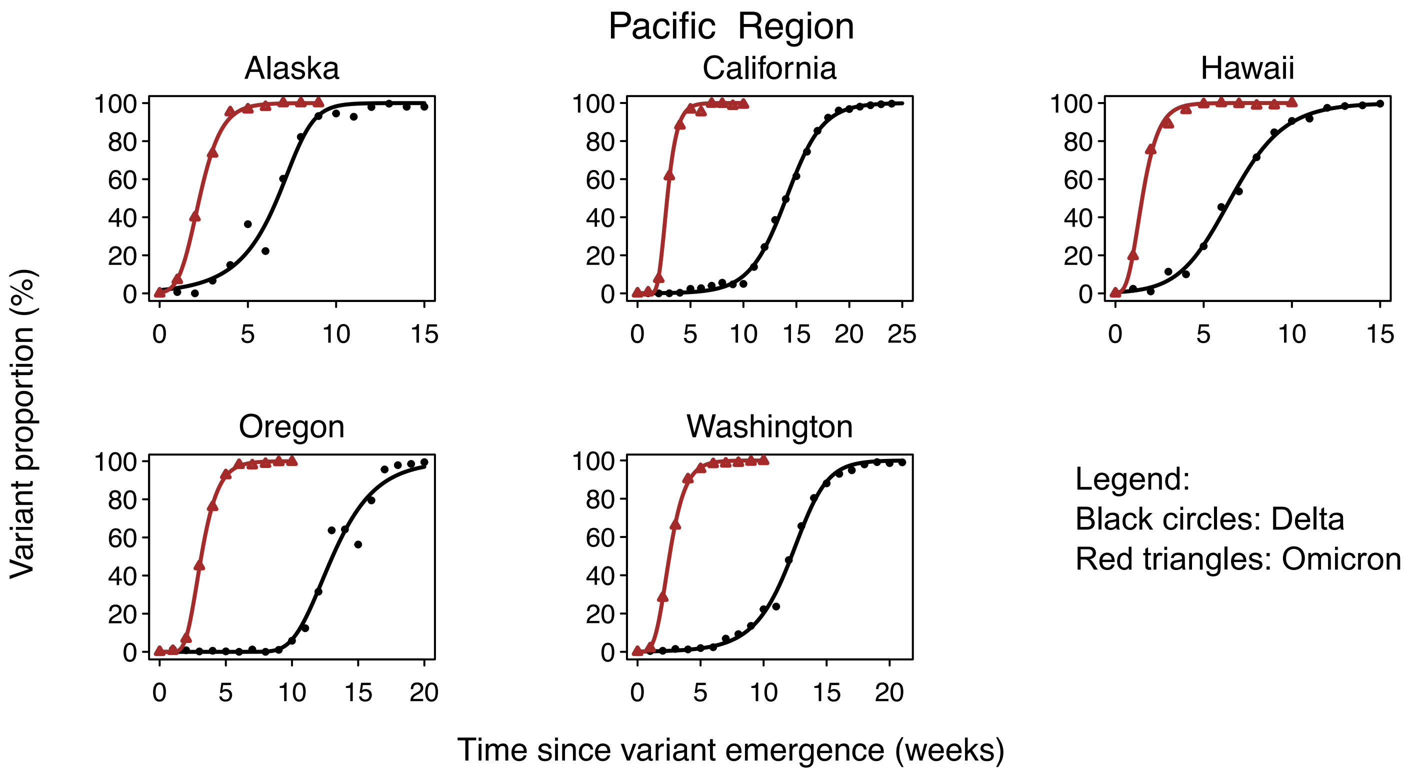

### Supplemental Figure 2

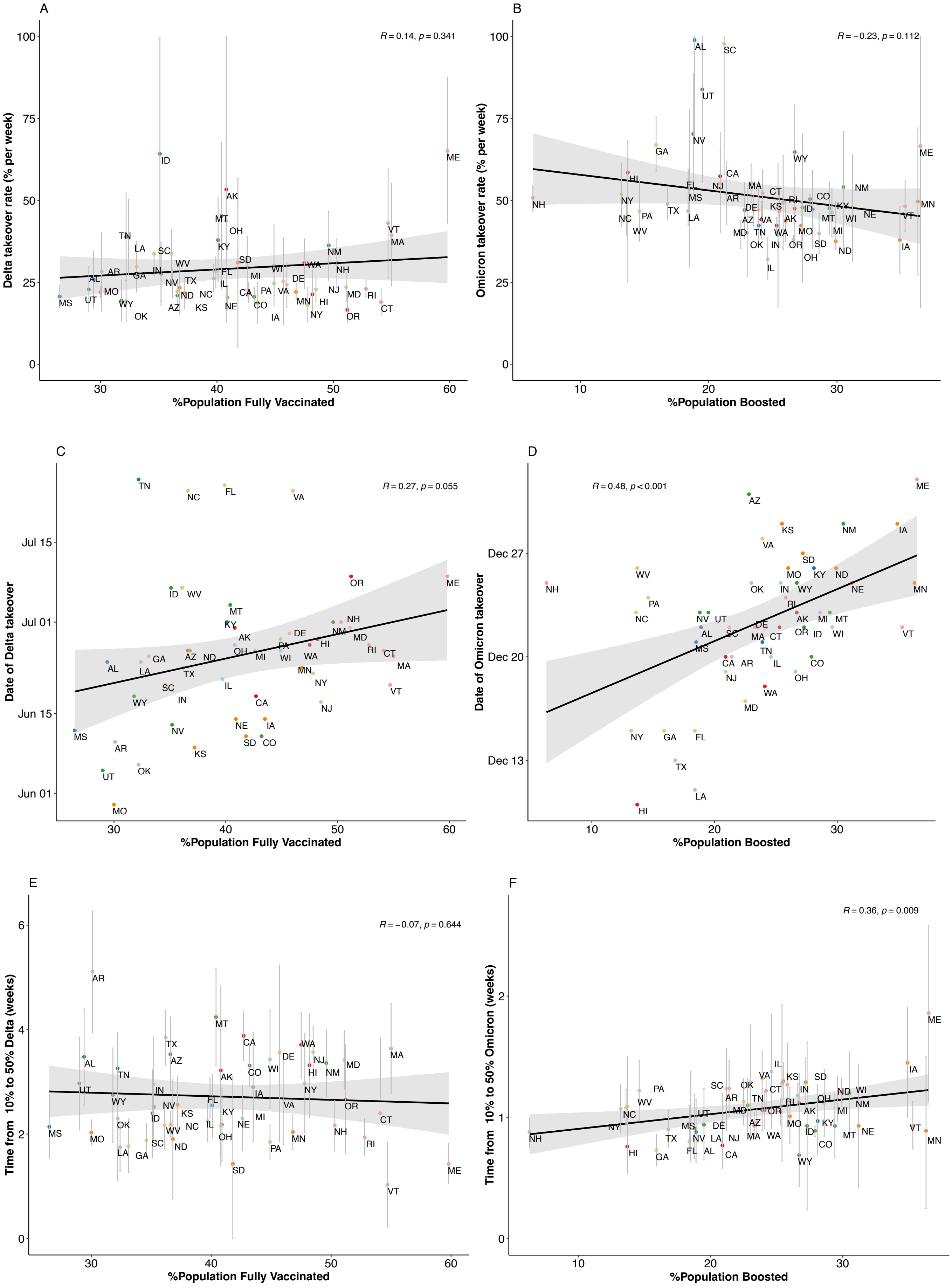

### Supplemental Figure 3

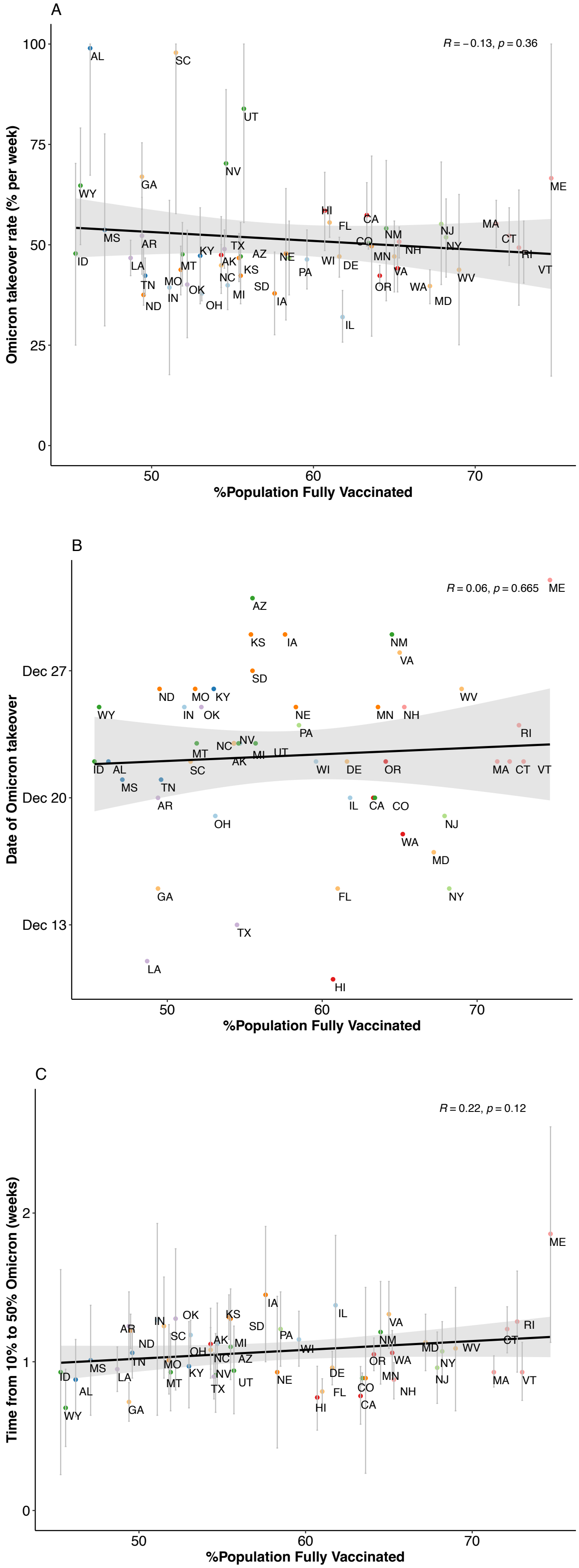

### Supplemental Figure 4

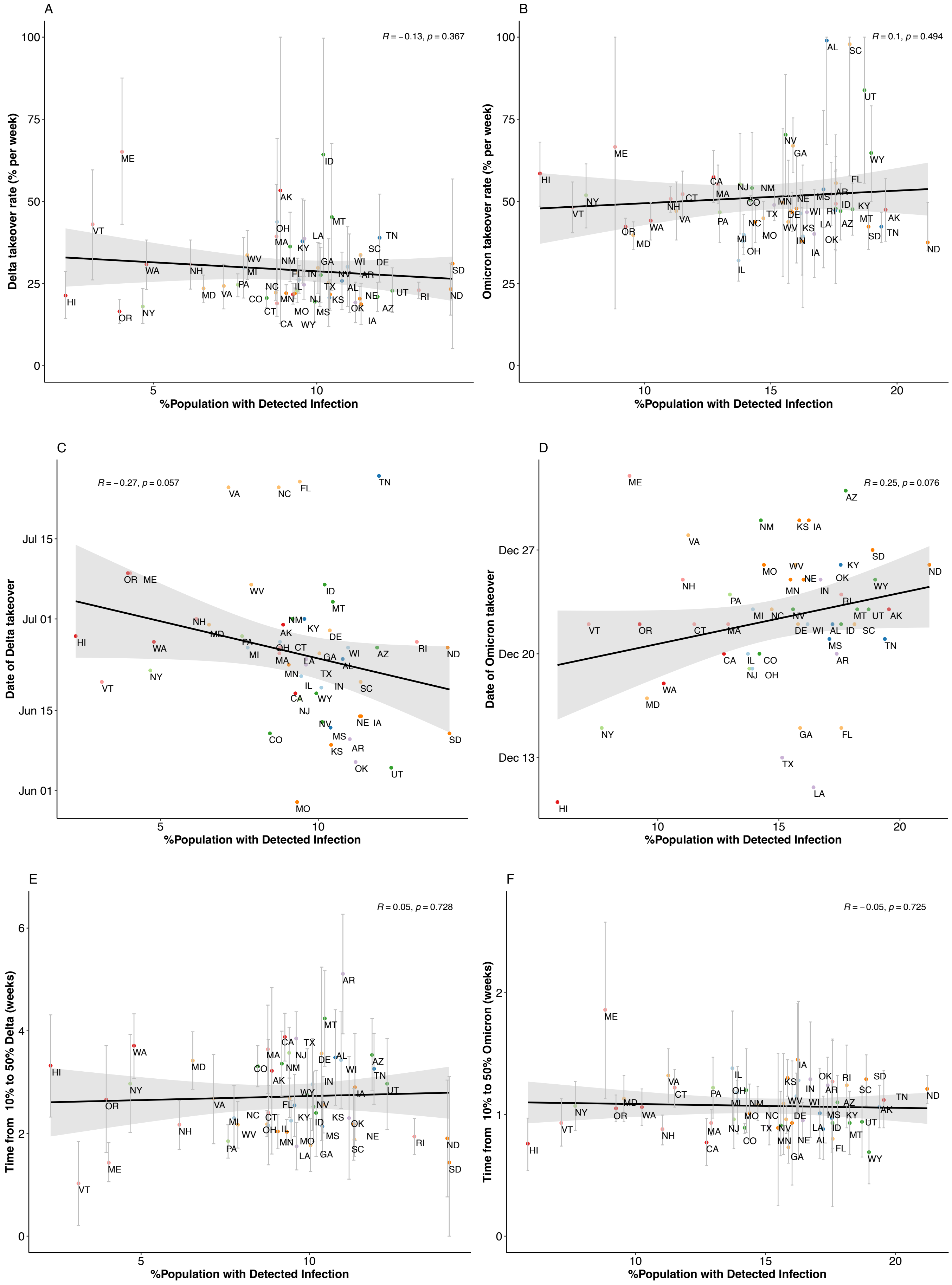
