## Supplemental Table 1 for "Pre-existing population immunity and SARS-CoV-2 variant establishment and dominance dynamics in the United States: An ecological study"

Table S1. Parameter estimates and standard errors for logistic curves fit to changing variant proportion in each state. States are grouped by census geographic region. Slope, time to 10% and time to 50% variant proportions are shown for Delta and Omicron.

|  | Delta | | | | | | Omicron | | | | | |
| --- | --- | --- | --- | --- | --- | --- | --- | --- | --- | --- | --- | --- |
| State | Slope (%Delta/week) | Standard error | Time to 10% (weeks) | Standard error | Time to 50% (weeks) | Standard error | Slope (%Omicron/week) | Standard error | Time to 10% (weeks) | Standard error | Time to 50% (weeks) | Standard error |
| **New England** | | | | | | | | | | | | |
| MA | 39.36 | 7.36 | 9.31 | 0.31 | 12.95 | 0.11 | 55.18 | 2.61 | 2.22 | 0.031 | 3.15 | 0.015 |
| CT | 18.98 | 1.88 | 7.708 | 0.218 | 10.11 | 0.109 | 52.24 | 3.05 | 1.95 | 0.046 | 3.17 | 0.021 |
| ME | 65.11 | 10.56 | 9.54 | 0.128 | 10.97 | 0.049 | 66.59 | 20.9 | 1.71 | 0.215 | 3.57 | 0.0927 |
| NH | 30.87 | 3.57 | 9.70 | 0.155 | 11.87 | 0.072 | 50.77 | 1.73 | 2.76 | 0.032 | 3.639 | 0.020 |
| RI | 22.99 | 1.18 | 8.37 | 0.105 | 10.31 | 0.061 | 49.3 | 6.25 | 2.18 | 0.0998 | 3.446 | 0.0479 |
| VT | 43.04 | 7.73 | 7.22 | 0.23 | 8.25 | 0.16 | 48.19 | 3.26 | 1.218 | 0.053 | 2.15 | 0.0288 |
| **Mid Atlantic** | | | | | | | | | | | | |
| NJ | 22.81 | 1.77 | 8.22 | 0.17 | 11.79 | 0.066 | 55.18 | 6.59 | 1.69 | 0.0696 | 2.65 | 0.035 |
| NY | 18.03 | 2.54 | 11.64 | 0.32 | 14.61 | 0.14 | 51.87 | 4.27 | 1.144 | 0.057 | 2.21 | 0.0275 |
| PA | 24.65 | 1.62 | 11.56 | 0.1 | 13.41 | 0.053 | 46.72 | 3.97 | 2.236 | 0.067 | 3.455 | 0.0327 |
| **East North Central** | | | | | | | | | | | | |
| IN | 27.59 | 4.6 | 9.21 | 0.309 | 12.17 | 0.118 | 39.36 | 9.2 | 1.428 | 0.177 | 2.71 | 0.095 |
| IL | 26.12 | 2.02 | 10.21 | 0.13 | 12.46 | 0.053 | 32.0 | 2.82 | 1.639 | 0.13 | 3.02 | 0.073 |
| MI | 29.80 | 4.41 | 8.88 | 0.21 | 11.18 | 0.091 | 39.91 | 2.58 | 2.28 | 0.077 | 3.396 | 0.0429 |
| OH | 43.78 | 11.78 | 8.17 | 0.27 | 10.33 | 0.097 | 37.89 | 0.77 | 1.587 | 0.025 | 2.77 | 0.0145 |
| WI | 30.09 | 5.74 | 7.66 | 0.317 | 11.09 | 0.129 | 46.35 | 3.2 | 2.005 | 0.0535 | 3.158 | 0.0253 |
| **West North Central** | | | | | | | | | | | | |
| IA | 18.73 | 2.94 | 4.51 | 0.334 | 7.41 | 0.157 | 37.89 | 4.49 | 2.755 | 0.140 | 4.21 | 0.0569 |
| KS | 21.70 | 2.13 | 5.19 | 0.173 | 7.75 | 0.084 | 46.72 | 2.5 | 1.879 | 0.044 | 3.179 | 0.021 |
| MN | 22.07 | 1.18 | 9.67 | 0.1 | 11.71 | 0.059 | 49.66 | 9.75 | 2.696 | 0.170 | 3.589 | 0.091 |
| MO | 22.00 | 0.77 | 4.20 | 0.072 | 6.23 | 0.043 | 43.78 | 2.61 | 2.698 | 0.0643 | 3.713 | 0.038 |
| NE | 20.42 | 1.73 | 4.24 | 0.20 | 6.42 | 0.125 | 47.82 | 6.88 | 1.729 | 0.135 | 2.66 | 0.081 |
| ND | 23.32 | 3.68 | 5.30 | 0.329 | 7.205 | 0.199 | 37.52 | 1.1 | 1.608 | 0.0308 | 2.815 | 0.0175 |
| SD | 31.05 | 11.77 | 1.537 | 0.525 | 2.97 | 0.242 | 42.31 | 3.08 | 1.53 | 0.0576 | 2.816 | 0.0309 |
| **South Atlantic** | | | | | | | | | | | | |
| DE | 25.31 | 6.29 | 6.054 | 0.58 | 9.609 | 0.214 | 47.09 | 2.09 | 1.21 | 0.0303 | 2.174 | 0.017 |
| FL | 28.33 | 2.21 | 9.71 | 0.13 | 12.4 | 0.052 | 55.55 | 1.64 | 1.216 | 0.0241 | 2.017 | 0.0127 |
| GA | 29.80 | 3.68 | 10.26 | 0.18 | 12.03 | 0.071 | 66.95 | 3.63 | 1.45 | 0.0427 | 2.18 | 0.0137 |
| MD | 23.54 | 2.06 | 10.32 | 0.19 | 13.74 | 0.076 | 39.73 | 1.78 | 1.329 | 0.0525 | 2.459 | 0.0293 |
| SC | 33.70 | 3.09 | 6.59 | 0.104 | 8.47 | 0.048 | 97.86 | 17.25 | 1.983 | 0.1075 | 3.215 | 0.0397 |
| NC | 22.26 | 1.36 | 10.11 | 0.078 | 12.29 | 0.041 | 44.88 | 2.35 | 2.266 | 0.0397 | 3.353 | 0.020 |
| VA | 33.70 | 3.24 | 9.54 | 0.31 | 12.22 | 0.11 | 47.09 | 3.83 | 1.804 | 0.0654 | 3.117 | 0.0327 |
| WV | 33.66 | 3.49 | 9.51 | 0.142 | 11.68 | 0.058 | 43.78 | 8.09 | 2.766 | 0.117 | 3.855 | 0.0616 |
| **East South Central** | | | | | | | | | | | | |
| AL | 25.83 | 4.16 | 7.3 | 0.319 | 10.78 | 0.12 | 98.96 | 13.69 | 2.217 | 0.0843 | 3.10 | 0.027 |
| KY | 37.89 | 6.14 | 10.31 | 0.21 | 12.86 | 0.078 | 47.27 | 5.11 | 1.705 | 0.0878 | 2.683 | 0.0373 |
| MS | 20.67 | 1.58 | 4.047 | 0.175 | 6.19 | 0.11 | 53.71 | 10.37 | 2.079 | 0.107 | 3.088 | 0.054 |
| TN | 38.92 | 6.40 | 9.31 | 0.238 | 12.57 | 0.089 | 42.31 | 1.99 | 1.98 | 0.0484 | 3.044 | 0.0279 |
| **West South Central** | | | | | | | | | | | | |
| AR | 28.29 | 5.7 | 3.71 | 0.407 | 8.82 | 0.146 | 52.24 | 4.0 | 1.621 | 0.0661 | 2.858 | 0.0292 |
| LA | 38.63 | 5.52 | 8.15 | 0.16 | 9.90 | 0.063 | 46.72 | 1.84 | 0.750 | 0.0401 | 1.699 | 0.0247 |
| OK | 19.24 | 2.94 | 7.02 | 0.362 | 9.32 | 0.208 | 40.1 | 5.81 | 2.21 | 0.141 | 3.50 | 0.0646 |
| TX | 24.65 | 2.13 | 8.81 | 0.18 | 12.66 | 0.071 | 48.93 | 2.09 | 1.005 | 0.0426 | 1.912 | 0.0263 |
| **Mountain** | | | | | | | | | | | | |
| AZ | 20.97 | 2.17 | 8.55 | 0.24 | 12.08 | 0.099 | 47.09 | 3.75 | 2.272 | 0.0621 | 3.371 | 0.032 |
| CO | 20.60 | 1.32 | 4.72 | 0.13 | 8.03 | 0.059 | 50.4 | 0.67 | 2.022 | 0.0114 | 2.908 | 0.0070 |
| ID | 64.23 | 16.92 | 12.36 | 0.314 | 14.76 | 0.090 | 47.82 | 9.79 | 2.25 | 0.195 | 3.179 | 0.103 |
| NM | 36.27 | 5.0 | 7.54 | 0.225 | 10.9 | 0.079 | 54.08 | 7.58 | 3.01 | 0.103 | 4.21 | 0.046 |
| MT | 45.25 | 10.26 | 2.11 | 0.321 | 6.35 | 0.107 | 47.64 | 3.18 | 0.309 | 0.0669 | 1.24 | 0.0397 |
| NV | 27.59 | 4.05 | 7.82 | 0.23 | 10.34 | 0.095 | 70.27 | 7.98 | 2.43 | 0.0769 | 3.34 | 0.0301 |
| UT | 22.81 | 3.31 | 6.12 | 0.31 | 9.09 | 0.11 | 83.88 | 12.14 | 2.41 | 0.09 | 3.35 | 0.035 |
| WY | 19.46 | 2.94 | 3.08 | 0.289 | 5.83 | 0.133 | 64.75 | 6.25 | 1.79 | 0.065 | 2.48 | 0.045 |
| **Pacific** | | | | | | | | | | | | |
| AK | 53.34 | 30.61 | 3.46 | 0.551 | 6.68 | 0.20 | 47.46 | 4.05 | 1.137 | 0.065 | 2.259 | 0.0337 |
| CA | 21.70 | 1.51 | 10.17 | 0.16 | 14.05 | 0.064 | 57.39 | 3.42 | 2.031 | 0.0519 | 2.803 | 0.032 |
| HI | 21.34 | 3.27 | 3.10 | 0.32 | 6.42 | 0.14 | 58.49 | 4.19 | 0.758 | 0.0557 | 1.518 | 0.036 |
| OR | 16.55 | 1.77 | 10.4 | 0.31 | 13.06 | 0.19 | 42.31 | 1.07 | 2.10 | 0.029 | 3.15 | 0.017 |
| WA | 30.90 | 3.57 | 8.51 | 0.22 | 12.22 | 0.078 | 44.15 | 2.46 | 1.47 | 0.044 | 2.53 | 0.023 |
